# Mapping the needs and psychological outcomes of Slovenian adolescent and young adult cancer patients: An exploratory mixed-methods study

**DOI:** 10.1101/2020.04.01.20049296

**Authors:** Urška Košir, Sanja Roškar, Jennifer Wild, Lucy Bowes

**Author notes:** **Corresponding Author:** Urška Košir, Department of Experimental Psychology, University of Oxford, Anna Watts Building, Radcliffe Observatory Quarter, Woodstock Rd, Oxford, OX2 6HG, United Kingdom.

## Abstract

**Objectives:** Baseline data from an ongoing online longitudinal survey were analysed. Three objectives guided our analyses: 1) to describe the self-reported levels of psychological distress among young Slovenian cancer patients and compare it to an international sample, 2) to describe the subjective illness experience of young Slovenian patients, and 3) to highlight areas for improvement in holistic care.

**Method:** Seventy-nine participants, aged 19 - 39 years, answered questionnaires about anxiety, depression, mental defeat, cancer worry, and their experience of learning the diagnosis and being treated. We used visualizations to demonstrate the relationship between anxiety and depression. The qualitative responses were analysed using an inductive thematic approach.

**Results:** Twenty-eight (35%) participants scored in the clinical range for anxiety and fifteen (19%) for depression. Cancer-related worry was common (85% reported at least one worry). Mental defeat was positively associated with measures of psychological distress. Those who felt negative about learning their diagnosis emphasized the need for more time, empathy, and dignity. Psychological support during illness was seen as crucial.

**Conclusion:** Anxiety and depression remain a problem for a subset of patients. Medical professionals working with young people with cancer should encourage a warm atmosphere as they attend to patients’ needs and concerns.

## Introduction

Despite the continuous and rapid progress in the development of novel therapeutic approaches in oncology, which result in ever increasing survival rates, there remain large discrepancies in outcomes between countries across Europe (Allemani et al., 2018). As the number of cancer survivors increases, so does the need to attend to the patients’ needs beyond treatment. A particularly vulnerable group of patients are adolescent and young adults (AYA), who have traditionally been overlooked, either placed together with children, or much older adults based on arbitrary age cut-offs, which vary between regions (Kosir, 2019). It has now been well established that the needs of AYA patients are different and unique to this age group (Barnett et al., 2016; Kosir, Wiedemann, Wild, & Bowes, 2019b; Sansom-Daly & Wakefield, 2013), and many efforts, as well as cross-border initiatives are promoted in hopes to empower young patients and shift the public and medical opinion, and even policy in order to adapt a more developmentally sensitive approach to care (Aldiss, Fern, Phillips, & Gibson, 2018; Buchacz et al., 2018).

Both Youth Cancer Europe (YCE), a leading pan-European organization for AYA patients, as well as UK’s James Lind Alliance emphasized the need for novel and tailored approaches to psychosocial care of young patients who face the diagnosis of cancer (Aldiss et al., 2018; Buchacz et al., 2018). Although psychological distress and anxiety are normal responses to life-threatening illness, many young patients continue to experience symptoms of distress upon reaching remission (Barnett et al., 2016; Kosir et al., 2019b; Sansom-Daly & Wakefield, 2013). The causes of such distress may be many and multifaceted; illness may impact cognitive processing and illness threat perceptions (Fardell et al., 2016; Rooij et al., 2018), cancer treatment may directly interfere with the endocrinological system (Jensen et al., 2018), illness can significantly derail achievement of normative milestones and disrupt education or an early career (H. M. Parsons et al., 2012; Vetsch et al., 2018), it can compromise family planning (Benedict, Shuk, & Ford, 2016; Levine, Canada, & Stern, 2010), cause a life-long disability (Barr, Ferrari, Ries, Whelan, & Bleyer, 2016), or significant financial strain (S. K. Parsons & Kumar, 2019).

Attending to the psychological needs of young patients is of vital importance as poorer psychosocial functioning can lead to missed medical appointments at outpatient clinics, and poorer quality of life (Klassen et al., 2015), which can in turn compromise the long-term outcomes. As poor mental health remains the leading cause of disability among young people (Gore et al., 2011), identifying the factors that contribute to psychological distress in young cancer patients will not only reduce the burden on individuals and their families, but also the burden on national health services.

Cancer is not a single illness and it does not only attack cells, but rather the entire person and their environment. The approach to treatment and care inevitably depends on the cultural context as well as on the health care system. Because survivorship is such a complex construct, developing novel psychosocial approaches must take into account one’s environment and culture. A more thorough understanding of young Slovenian patients’ needs is crucial for continued progress to improve outcomes on a national level, and it can highlight important factors that contribute to positive outcomes in AYAs, as well as areas of need that might have been overlooked abroad. Even though there exist well-established patient support groups in Slovenia, research activities are lagging behind, and to the best of our knowledge this is the first study of young Slovenian patients that aims to evaluate their needs and psychological well-being.

The objective of this study was 3-fold: 1) to describe the self-reported levels of psychological distress among Slovenian AYAs and compare it to an international sample of AYAs, 2) to describe the experience of young Slovenian patients from diagnosis through remission, and 3) to highlight areas on which nationally based strategies for improvement in holistic care could be based.

## Method

This study is based on baseline data from an ongoing longitudinal study exploring the predictors of psychological well-being and holistic recovery in AYA cancer patients. The study protocol has been registered and can be accessed at the following link: https://osf.io/d4e59/. This study has been approved by the Medical Sciences Inter-Divisional Research Ethics Committee at the University of Oxford (R61437/RE002).

Data were collected using Qualtrics, a secure online survey software provided by the university department, via an anonymous survey link distributed on social media and through patient advocacy groups. The survey has 2 equal arms, one that is fully translated into Slovenian, and the other, open to anyone across North America and Europe who is able to complete the questionnaires in English. Participants are adolescent and young adult cancer patients or survivors diagnosed with any form of malignant cancer. In the analysis, we included all Slovenian participants who completed at least 80% of the baseline questionnaires and were between 15 and 39 years of age at the time of their first diagnosis.

### Measures

### Demographic and Medical information

Participants reported their age, gender, relationship status, and level of education. They also reported information about their cancer and treatment type, age at diagnosis, and time since treatment completion.

### Stressful and Traumatic life events

In order to understand to what extent participants perceived their illness to be stressful or traumatic compared to other life events, they were asked to briefly describe up to 5 most stressful or traumatic life events and rank them in descending order.

### Patient Health Questionnaire (PHQ-9)

The PHQ-9 is a 9-item measure of depressive symptoms. Items are scored on a 4-point Likert scale from 0 (not at all) to 3 (nearly every day). A threshold score of 10 has been proposed to indicate the presence of clinical level of depressive symptoms (Kroenke, Spitzer, & Williams, 2001). The Cronbach Alpha for the Slovenian version of the questionnaire was 0.87.

### Generalized Anxiety Disorder (GAD-7)

The GAD-7 is a 7-item measure of anxiety symptoms (Spitzer, Kroenke, Williams, & Löwe, 2006). Items are scored on a 4-point Likert scale from 0 (not at all) to 3 (nearly every day). A sum score of 7 or above can indicate the presence of clinical anxiety (Plummer, Manea, Trepel, & McMillan, 2016). The Cronbach Alpha for the Slovenian version of the questionnaire was 0.89.

### Mental Defeat Questionnaire (MDQ)

The MDQ is an 11-item questionnaire, which asks about individuals’ perceptions of their mental state during a traumatic event (in this case, cancer illness) (Dunmore, Clark, & Ehlers, 2001). Agreement with statements such as *I lost any will power, I gave up, I felt like an object* are rated on a 5-point Likert scale from 1 (not at all) to 5 (very strongly). Higher total scores indicate greater presence of mental defeat during illness. The Cronbach Alpha for the Slovenian version of the questionnaire was 0.91.

### Cancer Related Worry

This 6-item Cancer Related Worry scale asks about the degree respondents agree with different worries related to cancer (e.g. relapse, fertility) on a 4-point Likert scale ranging from “strongly disagree” to “strongly agree”. The scale has been developed and validated in a Canadian AYA sample (Klassen et al., 2015). Only participants who were off-treatment completed this questionnaire. The Cronbach Alpha for the Slovenian version of the questionnaire was 0.75.

### Patients’ needs and experience

Individual patient’s experience was captured with 5 questions. A sliding scale from 0 to 100, where higher values represent better or more positive answers, was used to 1) report participants’ satisfaction with the way they received the diagnosis, 2) assess participants’ relationship with the medical team, and 3) describe the extent to which their needs as patients were met. The quantitative responses were supplemented with 2 open-ended questions that asked participants to briefly describe their experience with learning the diagnosis, and to describe what worked well during treatment, or else, what could have been improved. These answers yielded qualitative data.

### Analysis

We carried out exploratory analysis to visualize the relationship between anxiety and depression, and to compare scores of Slovenian participants to participants from abroad. Where relevant, we reported Spearman’s correlation coefficients. All analyses, data manipulations, and data visualizations were carried out using R Version 3.4.3.

Qualitative data were analysed using inductive thematic approach (Attride-Stirling, 2001; Elo & Kyngäs, 2008). Two researchers (UK and SR) independently read and coded all of the responses and rated individual experiences as either positive or negative. The researchers compared ratings of positive versus negative experience in diagnosis, and generated themes emerging from participants’ descriptions. Original themes were refined through discussion between the researchers and operationalized into a coherent list upon which all qualitative responses were mapped. Lastly, the classification of responses was compared and discrepancies resolved in a consensus meeting.

## Results Participants

Participants’ characteristics are presented in **Table 1**. Slovenian participants were significantly older (*M*= 33.7, *sd*=7.2) than English speaking responders (*M*=29.3, *sd*=5.75). Twenty-nine (37%) of Slovenian responders were in treatment at the time of the study, and the rest completed treatment on average 5.4 years earlier.

**Table 1:**
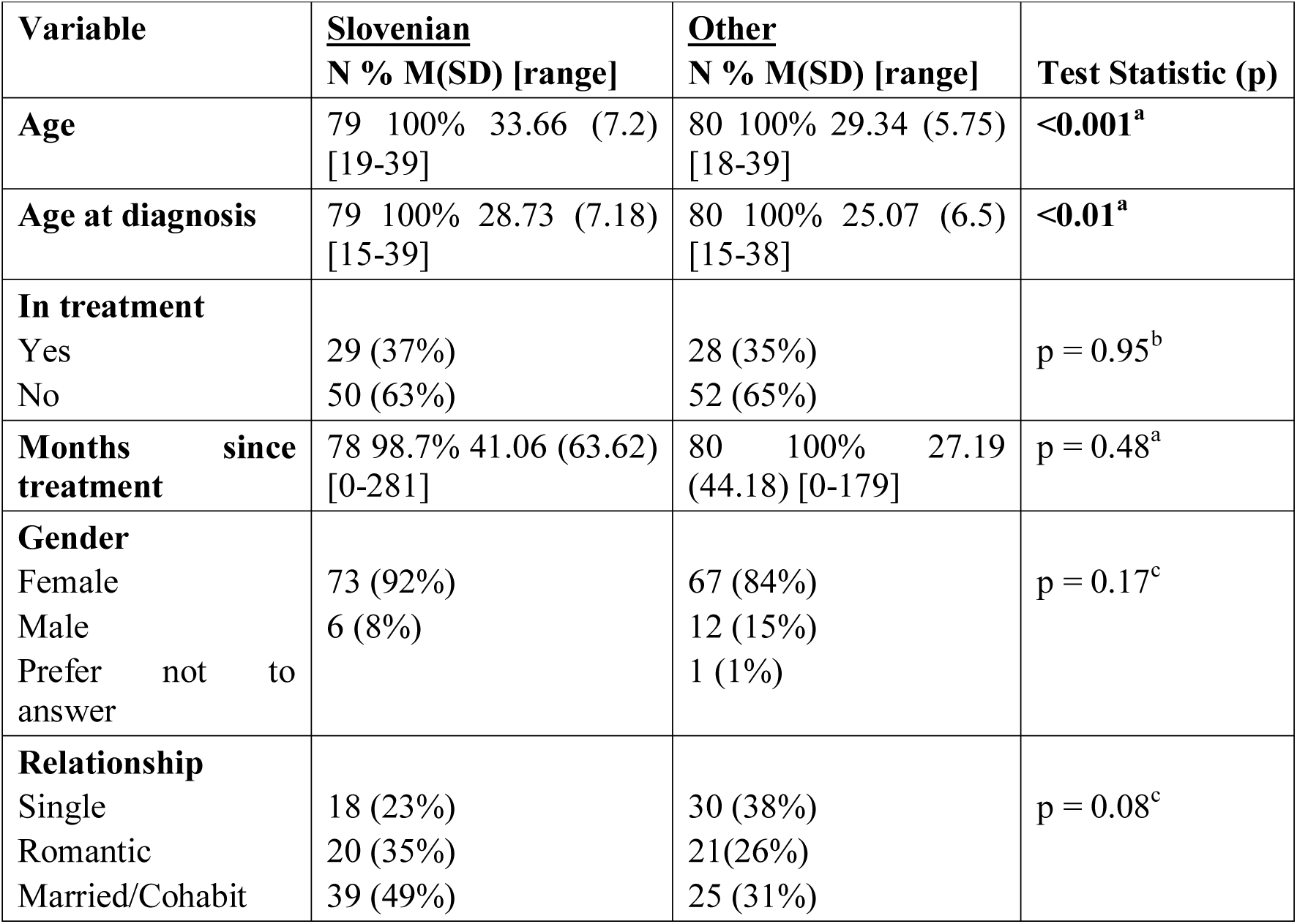
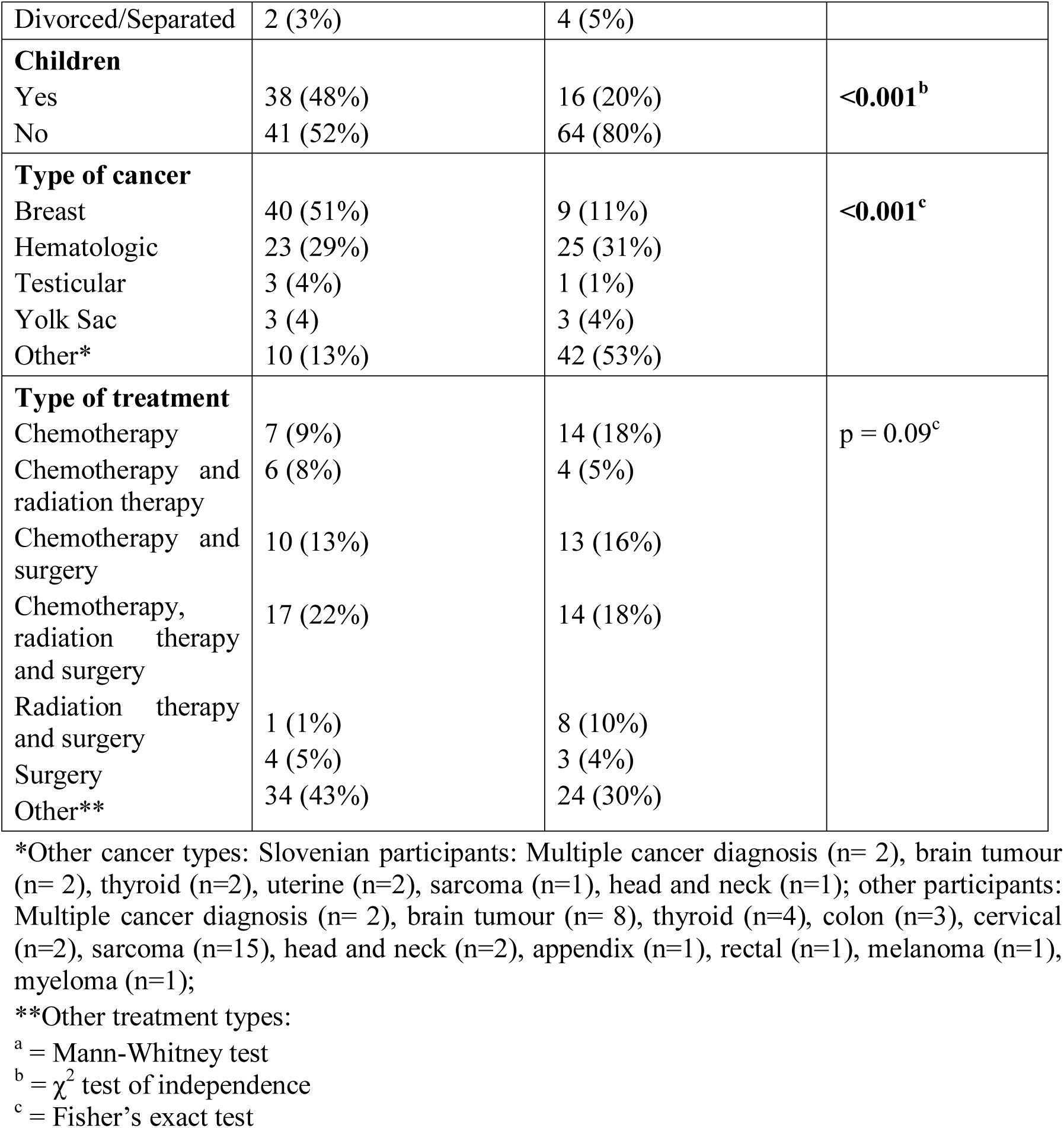
Demographic and Medical Information of participants

*Other cancer types: Slovenian participants: Multiple cancer diagnosis (n= 2), brain tumour (n= 2), thyroid (n=2), uterine (n=2), sarcoma (n=1), head and neck (n=1); other participants: Multiple cancer diagnosis (n= 2), brain tumour (n= 8), thyroid (n=4), colon (n=3), cervical (n=2), sarcoma (n=15), head and neck (n=2), appendix (n=1), rectal (n=1), melanoma (n=1), myeloma (n=1);

### Psychological well-being

Participants completed questionnaires about depression, anxiety, mental defeat, cancer worry, and stressful and traumatic life events. Scores of depression and anxiety are visually represented in **Figure 1** and plotted against data from the English arm of the same study. Similar proportion of Slovenian and other patients were in treatment at the time of the study (see **Table 1**). Higher scores in depressive symptoms were correlated with higher scores in anxiety in both groups (Spearman’s rho; *r* = 0.78, *p* <0.001). As a group, Slovenian patients report lower scores of psychological distress, however, they are older (see **Table 1**). **Table 2** shows average scores on the self-reported measures for the Slovenian population. Seventy-six participants reported having stressful or traumatic life events, and among those, 53 (70%) reported cancer to be among the three most stressful or traumatic events in their lifetime. Mental defeat was also positively correlated with anxiety (Spearman’s rho; *r* = 0.4, *p* <0.001), and depression (Spearman’s rho; *r* = 0.41, *p* <0.001).

**Table 2:** Mean scores on measures of psychological distress

|  | **N, mean (SD) [range]** |
| --- | --- |
| **Anxiety** | 79, 5.7 (4.86) [0-17] |
| **Depression** | 78, 5.27 (5.0) [0-20] |
| **Mental Defeat** | 76, 7.22 (7.69) [0-37] |
| **Cancer worry\*** | 47, 7.64 (3.7) [0-16] |
|  | **N (%)** |
| **Cancer as trauma** |  |
| Yes | 53 (69.7) |
| No | 23 (30.3) |
| **Satisfaction with receiving diagnosis** | 76, 63.71 (37.34) [0-100] |
| **Relationship with the medical team** | 76, 88.39 (18.99) [0-100] |
| **Extent of needs met** | 76, 80.18 (22.36) [5-100] |

\*Cancer worry questionnaire was only administered to participants who completed their treatment in order to reduce bias and inflating worry. Questions ask if a person thinks about cancer every day, which might be normal in the acute phase of cancer treatment.

**Figure 1:**
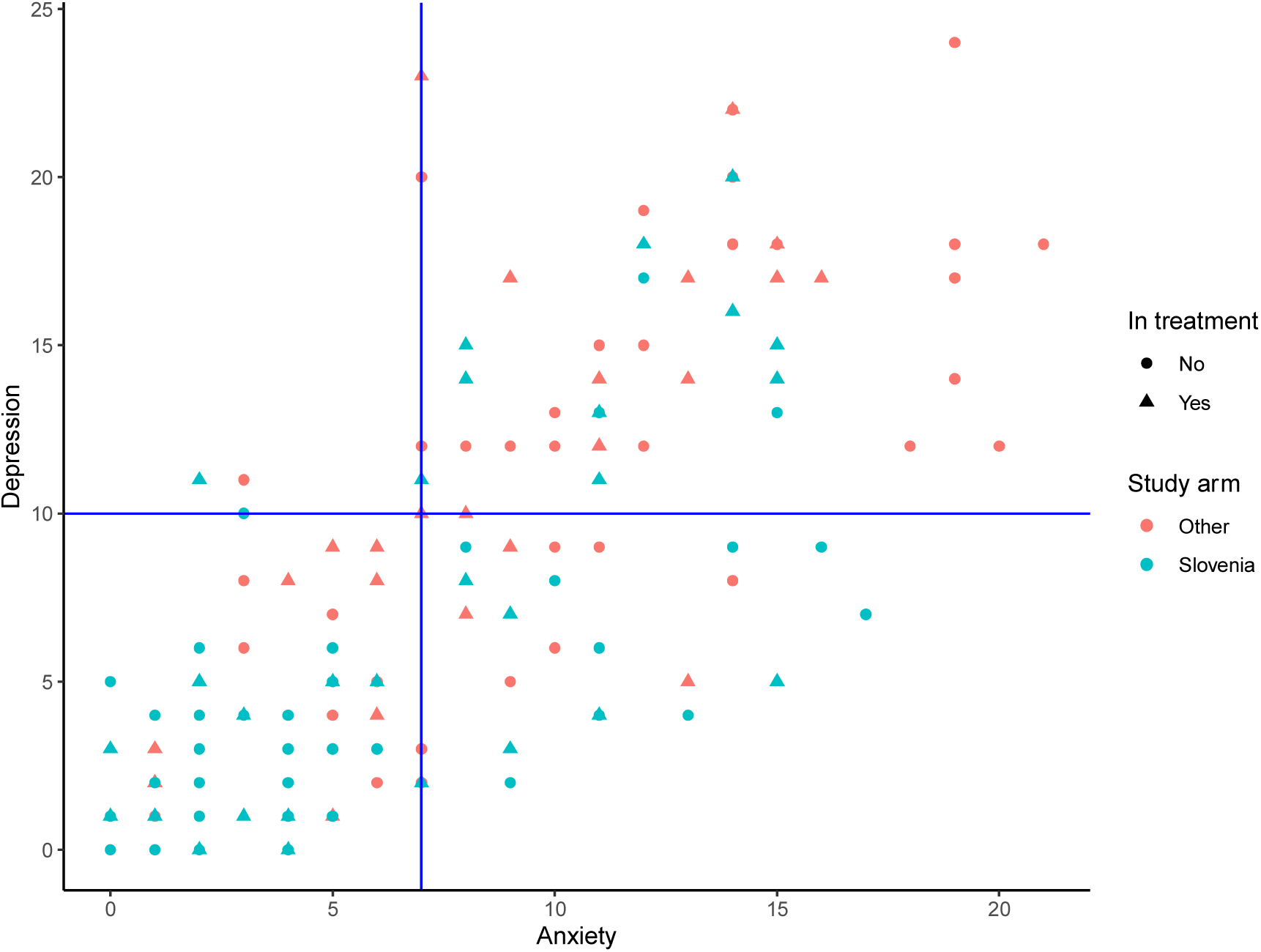
Scatterplot showing the relationship between anxiety and depression in Slovenia and abroad. Different colours represent different regions, while different shapes represent patients who were in or off-treatment at the time of the study. The blue lines on the graph represent the clinical cut-off levels for anxiety and depression, respectively. Participants labelled as Other were young patients and survivors who were able to respond in English. Most participants were from the UK, Sweden, Canada, USA, and Netherlands. Several other European nationals participated. Twenty-eight (35%), and 15 (19%) Slovenian participants scored in the clinical range for anxiety and depression, respectively. Of the former, 16 were in treatment, while 11 of those who scored in the clinical range for depression were in treatment at the time of the study.

## Experience

### Learning the diagnosis

Seventy-seven (97%) participants responded to an open-ended question about the way they learned their diagnosis. Three response groups emerged; the majority (55%) reported the experience to be positive, some were ambivalent (6%), and many (39%) reported a negative experience.

Among the five most commonly reported factors endorsed by those with a positive experience in learning their diagnosis were: experiencing a positive, encouraging and supportive atmosphere within the medical team, empathy, professional approach, adequate explanation and information provided, and having a clearly outlined treatment plan.

> *“Ever since I found out about the diagnosis, I was very pleased with the treatment and attitude of the doctors and nurses. My oncologist took the time (over an hour) to explain the course of treatment and answer all of my and my husband’s questions*.*”* (Female, 39)

Among the top five reported factors that contributed to a negative experience in learning the diagnosis were: being rushed or having inadequate time with the medical team, lack of empathy, lack of dignity, minimizing or invalidating one’s experience with cancer, and being given an overwhelming amount of information at once.

> *“I received my diagnosis on the hallway of the clinic. The hallway was full of people. The doctor called me and told me the result of the puncture. She was with me for 2 minutes, then she left. In consolation, she said that I would not die from this cancer*.*”* (Female, 29)

Those who were ambivalent reported that their individual experience was not optimal; one person reported inappropriate use of humour by a medical professional, while another person reported that they did not appreciate that their parents were told before them. **Figure 2** demonstrates emergent themes.

**Figure 2:**
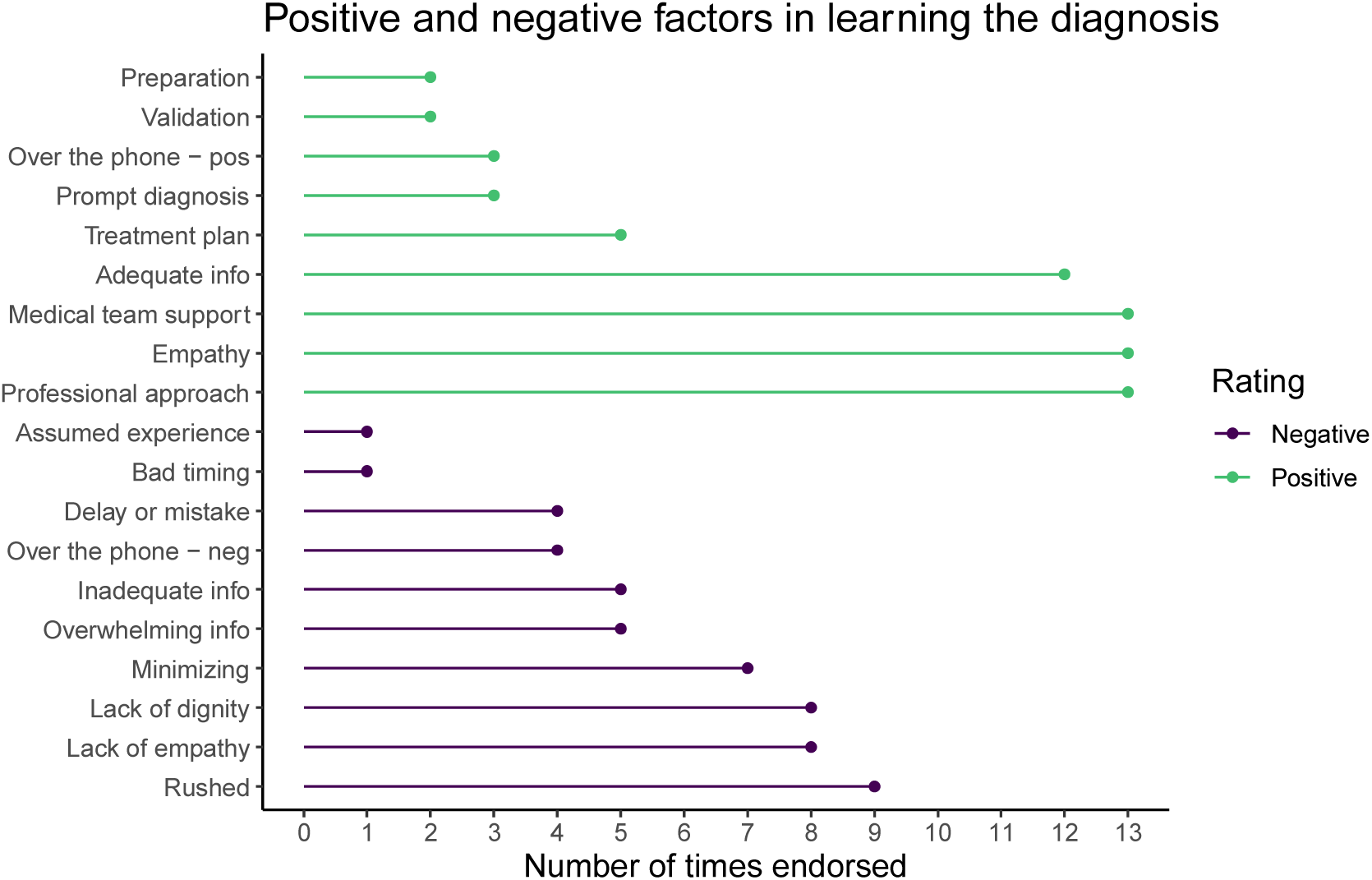
Positive and negative factors in learning the diagnosis. The figure demonstrates the number of times a specific factor, either positive or negative, contributing in the overall experience in learning the diagnosis was endorsed in short answers. *Adequate info* = participants report being happy with the amount of relevant information about their disease and treatment; *Treatment Plan* = Having a clear plan and steps ahead; *Over the phone – pos/neg* = learning the diagnosis over the phone was positive for some while negative for others; *Preparation* = feeling prepared for the bad news; *Validation* = validation of emotional distress; *Assumed experience* = because of medical background medical doctors assumed the patients was well informed about the disease; *Delay or mistake* = lag in diagnosis or misdiagnosis; *Minimizing* = telling patients they have a “good cancer”; *Rushed* = feeling rushed through the diagnosis.

**Figure 3:**
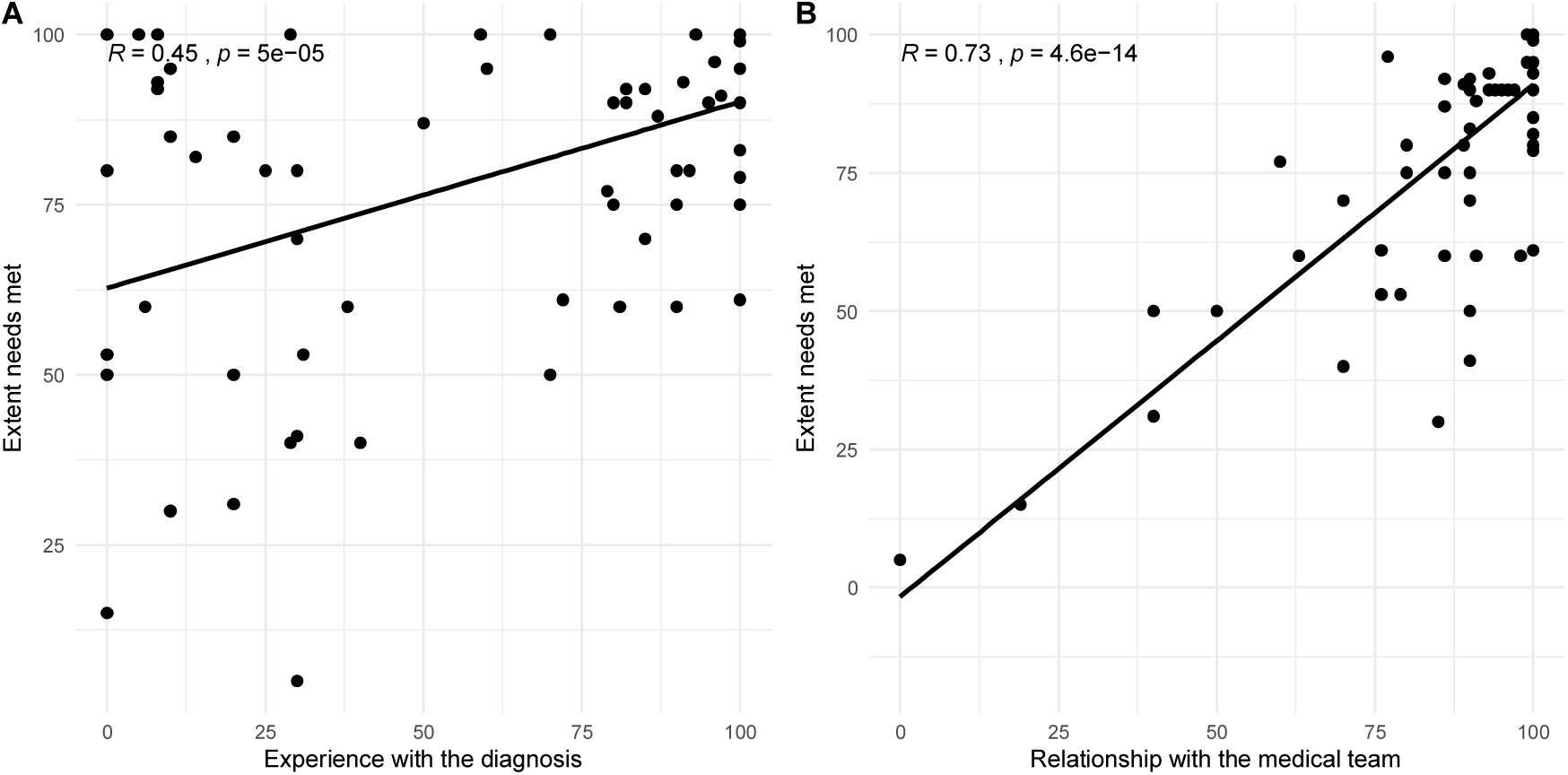
Scatterplots showing association between the **A**) self-rated experience in learning the diagnosis and **B**) relationship with the medical team (higher values indicate more positive experience) and the self-reported extent of patient’s needs met (higher value indicates more needs met). The correlations reported are Spearman’s Rho coefficients, and the grey area around the line represents standard error.

### Patients’ needs

Participants were asked to estimate the extent to which their needs as young patients were met, as well as to provide a short descriptive answer about their experience to support their response. The ratings on the numeric scale were skewed to the left, with the majority reporting that their needs were met to a great extent (see **Table 2**).

Sixty-nine participants provided short answers describing their experience, what worked, who offered most support and what could have been done better. Among those who made reference to support, 18 endorsed their medical team as a great source of support, 14 emphasized the importance of their family, and 10 drew support from their close friends.

Participants provided varying degrees of detail in their answers about what helped them most during their illness. Beyond the already mentioned supportive framework within the medical settings, they described many ways in which they coped. Three most commonly endorsed aspects that most helped patients through their illness were: receiving psychological support, participating in a patient support group (be it online or in person), and feeling heard and listened to, especially among the medical professionals. Other ways of coping to which patients made explicit references are described in **Table 3**.

**Table 3:**
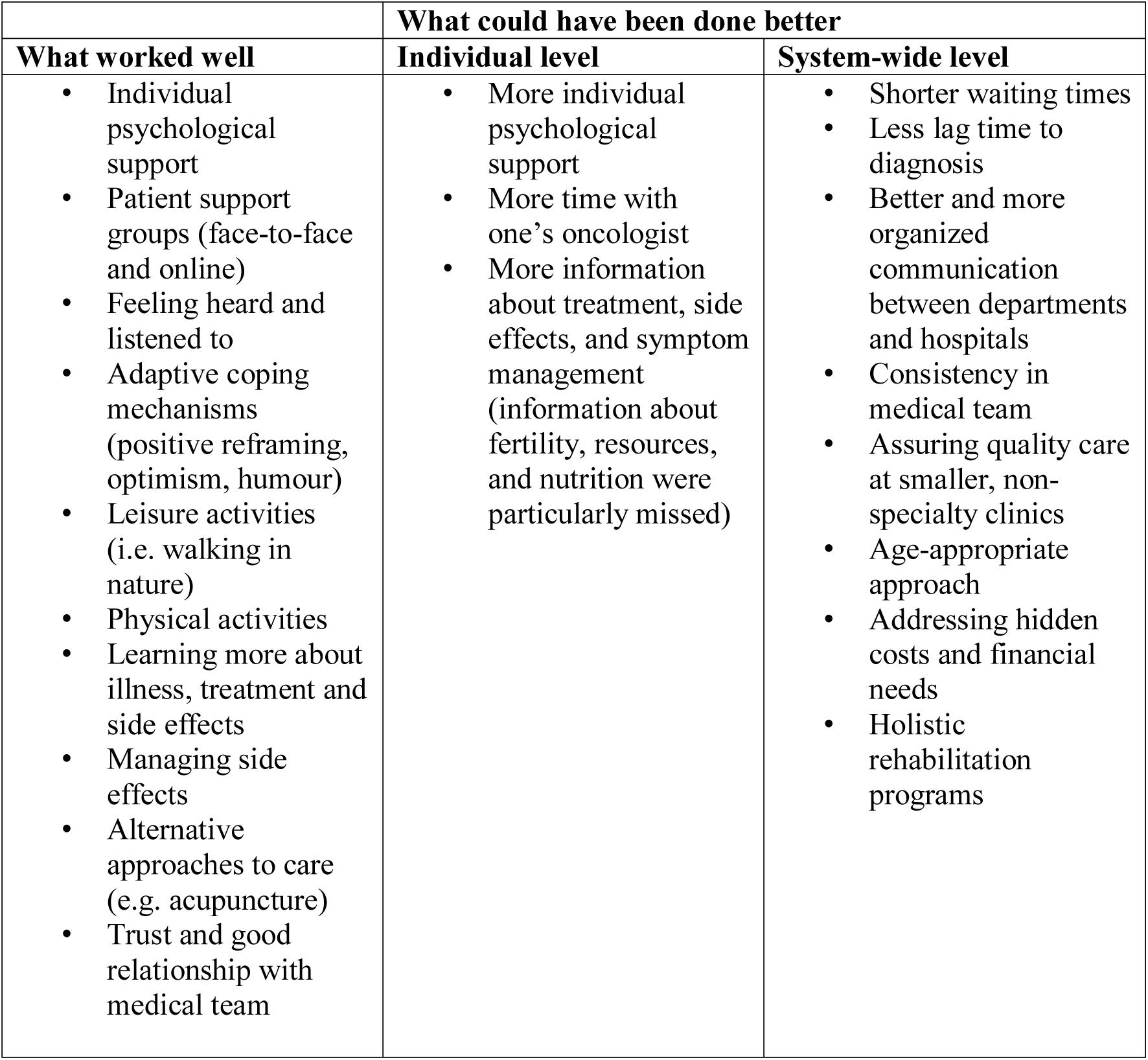
Themes emerging from participants’ account of illness experience describing what worked well and what could be improved

The majority of the patients also provided answers about what they thought could have been done better. These could be divided into personal experience and system-wide changes. In the former, the majority reported the need for a structured psychological evaluation and intervention, followed by the need to have more time with their primary oncologist, especially during the first consultation, and a more age-appropriate approach, services, and resources.

> *“There is lack of guidance and tips on how to cope with cancer diagnosis on a psychological level*.*”* (Male, 31)

Several patients also noted that there was a significant delay in their diagnosis due to long waiting times, and reported frustration with lack of, or poor communication between different departments (see **Table 3**).

## Discussion

This is the first study to describe the psychological well-being and experience of cancer in a subset of young Slovenian cancer patients. The analyses were guided by three main objectives: to assess the level of psychological distress among Slovenian AYA patients and to compare it to an international sample, to describe the experience of learning the diagnosis and to provide directions for national service and policy development.

Mental health outcomes and psychological well-being are increasingly considered and evaluated in young cancers patients (Aldiss et al., 2018; Barakat, Galtieri, Szalda, & Schwartz, 2016; Barr et al., 2016; De et al., 2019). However, studies continuously yield conflicting results, which might in part be attributable to a lack of psychological measures adapted for cancer populations. Our analysis showed that symptoms of anxiety and depression often co-occur, and that regardless of treatment status (in- or off-treatment) there remains a subset of individuals who score in the clinical range for mood disorders based on self-report assessment. As a group, young Slovenian patients scored lower on both measures of psychological distress compared to the international sample. However, it is important to note that the international sample was significantly younger, and not matched on demographic or illness characteristics.

Mental defeat, which has been proposed as a potential cognitive mechanism contributing to psychological distress in a pilot phase of this study (Kosir, Wiedemann, Wild, & Bowes, 2019a) shows strong association with both anxiety and depression, however due to the cross-sectional nature we did not assess the causal mechanisms.

As previously reported (Kosir et al., 2019a; Wang et al., 2015), cancer-related worries remain highly prevalent among the AYAs, with 85% worrying about at least one aspect of their cancer illness. Interestingly, even though worry has been identified as a predictor of anxiety and even depression (Muris, Roelofs, Rassin, Franken, & Mayer, 2005; Segerstrom, Tsao, Alden, & Craske, 2000), the association between cancer-related worry and symptoms of anxiety and depression in our study remains weak. Such symptoms may be exacerbated by having a cancer diagnosis rather than the outcome of a diagnosis. Future research will need to tease apart the relationship between cancer worry and symptoms of anxiety and depression in AYAs.

The majority of young cancer patients have little experience in navigating medical systems yet have distinct preferences for their cancer care (Pannier et al., 2019). The idea that the initial experience can have an impact on the way one interacts with the medical team and can influence the rest of the experience is not far-fetched. Our results reveal that those reporting a more positive experience at diagnosis, also reported having their needs met, as well as a better relationship with their medical team. A better relationship with the medical team was also associated with individuals’ reports of having their needs met. This may be because the initial experience leads to patients’ being more comfortable in voicing their concerns and needs, and results in a better, more patient-cantered approach to care.

Just over half of the participants reported a good experience in learning their diagnosis. The majority of those reported the importance of an empathic approach, receiving adequate information and explanation, feeling supported by the medical team, and having appreciated the professional attitude of medical workers. On the other hand, those with a negative experience reported feeling rushed and not having adequate time for all their questions, reported being treated with a lack of empathy and dignity, and described that the medical professionals minimized their experience either by underscoring the impact of a diagnosis, labeling it as “an easy cancer” or initially not believing their symptoms due to the young person’s age. Delay in cancer diagnosis in young people has been observed previously and partly attributed to failure in recognizing symptoms or signs of the disease, as well as unique biology of disease in AYA populations (Barr, 2014; Barr et al., 2016).

Additionally, several patients underwent multiple procedures because of inconclusive initial biopsies. This often resulted in delays in receiving their diagnosis, and increased uncertainty and frustration among young patients. A few individuals who reported a negative overall experience with the way they received the diagnosis further emphasized that once they were relocated or met with a different medical team, what worked better was being treated with more dignity and developing a concrete treatment plan. These themes seem to be common in contributing to a better experience of learning the diagnosis and allowing patients to feel at ease, empowered and relieved, knowing what they can expect next.

Despite the fact that in Slovenia it is not recommended to disclose a diagnosis to a patient over the phone, such has been the case for seven participants. Interestingly, receiving the diagnosis via a telephone call has been considered both a positive and a negative experience. The former was the case, when the patients requested to learn the diagnosis in what they referred to as a more convenient way which also saved time otherwise lost to commuting to a follow-up appointment. Hearing the diagnosis over the phone was negative in cases when patients were not expecting to receive any call and would have preferred being told in person. This example illustrates the importance of individual differences and points at an important aspect to be discussed earlier in the initial phases when cancer is first suspected.

Because cancer impacts multiple domains in one’s life, particularly at a younger age (e.g. school or vocational disruption, family planning) understanding the overall experience and what helps patients to cope best and what could be improved carries important implications for the development of age-appropriate services. A recent report by All.Can (All.Can, 2019), an international multi-stakeholder initiative to optimize policy in cancer care, which surveyed almost 4,000 adult patients from 10 countries across Europe (Slovenia was not included) described four main areas to be targeted for improvement of efficiency in cancer care. These were: 1) swifter diagnosis (including more empathy), 2) more information and the need for shared decision-making, 3) better coordinated approach and the need for integrated and multidisciplinary care, and 4) lessening the financial burden of cancer. Our study highlights the importance of the first three aspects in the approach to cancer care, and further calls for a developmentally sensitive, age-appropriate approach. The financial aspect seldom appeared in the responses of Slovenian participants. However, we did not explicitly ask about the financial impact of their illness. Nevertheless, three individuals made reference to the financial aspect of illness in their answers, while several others mentioned resorting to private care, paying out of pocket in order to speed up the diagnostic process. There may be substantial hidden costs to cancer care and treatment in Slovenia, even with a nationally based health care system.

## Limitations and future directions

This is the first study addressing the needs and outcomes of AYAs in Slovenia. Despite valuable insights, certain limitations should be taken into account. The study is based on a small subset of young Slovenian patients, which only included six male participants. This may in part be due to cultural factors that mean men are less likely to disclose or talk about emotions. Optimizing ways of reaching young men should be explored. Moreover, the online nature of the study implies a self-selected group of participants. Recall bias will influence the qualitative responses to the way patients received their diagnosis. Moreover, participants’ current psychological state might have influenced answers to the open-ended questions. As such, generalizations should be made with caution.

Slovenian participants were compared to an international sample from the same study. The participants were not matched, nor do they stem from the same socio-cultural environment, which may be pertinent in considering the psychological effect of cancer diagnosis and treatment. Future studies and analysis should aim at including a matched healthy population control in order to get a better sense of the psychological functioning of young Slovenian cancer patients relative to the Slovenian context. Lastly, most measures reported do not have an established construct validity, or have been fully validated in the Slovenian context. A comprehensive validation study of the Slovenian measures would benefit future research.

Additionally, half of the Slovenian sample had a diagnosis of breast cancer, which further limits the generalizability of our findings. This may also be due to the fact that EuropaDonna, a pan-European breast cancer organization for women’s empowerment has among the longest and strongest presence in Slovenia, with a subsection for young patients. This suggests that establishing more type- and age-specific patient organizations could lead to increased empowerment and greater research participation in the future.

The findings presented in this article are based on data from the baseline of a larger study, which makes this research cross-sectional in nature. In order to gain a better understanding of how mental defeat influences psychological distress or cancer-related worries, longitudinal data are needed.

### Clinical implications

This study is the first study to conduct an in-depth analysis of Slovenian AYA patients’ psychological functioning and needs. Our results show that a subset of Slovenian patients experience clinically elevated symptomatology of anxiety and depression. Targeting mental defeat early and during cancer treatment might alleviate anxiety and depression. Participants who reported receiving psychological support during their illness fared better and those who have not received psychological support, reported a need for such support. Development of novel and age-appropriate services and resources, including patient support groups should become a national priority. Additionally, medical professionals working with this population should aim to promote a warm and encouraging atmosphere, assure that patients feel heard, and provide adequate information and explanation to help guide patients and their relatives through the initial phases of cancer illness.

## Data Availability

Data are not available.

## Acknowledgment

Authors would like to thank all participants who took the time to provide thoughtful responses about their illness experience. First author, Urška Košir would also like to acknowledge the financial support for her studies from Economic and Social Research Council (ESRC) as well as Ad Futura by Public scholarship, development, disability and maintenance fund of the Republic of Slovenia. Moreover, thanks to the following organisations in Slovenia: EuropaDonna, Lymphoma and Leukemia Society, OnkoMan, and OnkoNet for help with recruitment and continued support of research endeavours.

